# Dissecting the comprehensive relationship between blood pressure and bone mass: the observational association and pleiotropic drug targets

**DOI:** 10.64898/2026.08.17.26360637

**Authors:** Jia-Xuan Gu, Meng-Yuan Yang, Xin Li, Peng Wei, Zeng-Hui Gu, Ming-Yu Han, Jia-Sheng Yu, Wang-Jun Chen, Zheng-Rui Liao, Si-Rui Gai, Jia-Dong Zhong, Pian-Pian Zhao, Bo Zhang, Zhi-Hai Fan, Ching-Lung Cheung, David Karasik, Hou-Feng Zheng

## Abstract

**Background:** Hypertension is a major global health challenge with well-established cardiovascular risks, yet its relationship with bone mineral density and the skeletal relevance of antihypertensive-related targets remain unclear.

**Methods:** Based on individual-level data from 366,443 European-ancestry participants in the UK Biobank, this study adopted restricted cubic spline models to explore linear and nonlinear associations between systolic/diastolic blood pressure (SBP/DBP) and heel estimated bone mineral density (BMD). We stratified participants by median DBP to conduct systematic biomarker analyses covering renal, endocrine, inflammatory and metabolic indicators. Drug-target Mendelian randomization (MR) combined with colocalization and mediation analyses was further performed to identify and validate causal antihypertensive-related target genes associated with BMD.

**Results:** A significant inverted U-shaped association was identified between DBP and BMD (P-nonlinear=3.23×10^-9^), with peak BMD observed at a DBP of 80–90 mmHg, while SBP showed a trend of nonlinear correlation. Biomarker analyses revealed that renal biomarker cystatin C and endocrine biomarker IGF-1 exhibited DBP-dependent associations with BMD, mediating the nonlinear DBP-bone density relationship. Drug-target MR demonstrated that genetically proxied *MMP9* expression (ACE inhibitor-related) was negatively correlated with BMD (β=-0.036, P=2.59×10^-6^), whereas *CACNA1G* expression (T-type calcium channel blocker target) was positively associated with BMD (β=0.042, P=1.57×10^-9^).

**Conclusion:** The inverted U-shaped association between blood pressure and bone mass might partly reflected by renal dysfunction. Antihypertensive pathways mediated by *MMP9* and *CACNA1G* exert opposing effects on bone mass, implying that skeletal health should be considered when selecting antihypertensive agents for vulnerable older populations.

**Novelty and Relevance:** 

**What Is New?:** This large-scale population study identifies a significant inverted U-shaped association between diastolic blood pressure (DBP) and bone mineral density (BMD), with an optimal DBP range of 80–90 mmHg for preserving bone mass. The study reveals that renal biomarkers (cystatin C and IGF-1) exhibit DBP-dependent variation patterns that underpin the nonlinear blood pressure–bone density linkage. Using systematic drug-target Mendelian randomization and colocalization analyses, we further prioritize *MMP9* and *CACNA1G* as antihypertensive-related targets associated with bone mass.

**What Is Relevant?:** This study demonstrates a nonlinear association and clarifies biomarker-mediated renal mechanisms linking blood pressure homeostasis to bone metabolism. The drug-target MR design minimizes confounding and provides reliable genetic evidence to explain why different antihypertensive pathways exert distinct skeletal outcomes.

**Clinical/Pathophysiological Implications?:** The optimal DBP range for bone preservation is 80–90 mmHg. Both excessively low and elevated DBP disrupt cystatin C and IGF-1 homeostasis, ultimately impairing bone formation and mineralization. Differential skeletal effects of *MMP9*-related ACE inhibitor pathways and *CACNA1G*-related T-type calcium channel pathways suggest the need for individualized antihypertensive strategies.

## Introduction

As populations age, hypertension has become a prevalent health concern among older adults, significantly increasing the risk of cardiovascular events and premature mortality^1–3^. Globally, around 1.13 billion individuals are affected by hypertension, with two-thirds residing in low- and middle-income countries^4^. Public health interventions and antihypertensive medications have improved blood pressure control, and pharmacological treatment remains a central component of hypertension management^5,6^.

Beyond its cardiovascular consequences, hypertension may also be relevant to skeletal health, particularly in relation to bone mineral density (BMD), although the direction and mechanisms of this association remain uncertain. Potential pathways include increased calcium excretion, heightened sympathetic nervous system activity, chronic inflammation, altered parathyroid hormone regulation, and renal dysfunction^7–10^. However, epidemiological evidence linking blood pressure to BMD remains inconsistent. Hypertensive women have been reported to exhibit lower bone density and higher urinary calcium excretion than normotensive women^11^. Conversely, other population-based cross-sectional studies have reported positive associations.

Specifically, analyses from the US NHANES cohort^12^ and the Canadian Multicentre Osteoporosis Study (CaMos)^13^ revealed that hypertension was associated with significantly higher BMD. These discrepancies may partly reflect population differences, residual confounding, limited power in continuous blood pressure analyses, and insufficient assessment of nonlinearity. Large-scale studies are therefore needed to clarify whether blood pressure components show nonlinear associations with BMD and whether these patterns differ between systolic and diastolic blood pressure.

Pharmacological treatment is central to hypertension management, particularly among individuals with sustained hypertension or elevated cardiovascular risk^14–16^. Beyond cardiovascular outcomes, some studies have indicated that antihypertensive medications may influence the risk of certain non-cardiovascular diseases, such as chronic kidney disease^17,18^, type 2 diabetes^19^, and Alzheimer’s disease^20^. Over the past decade, there has been growing debate regarding the relationship between antihypertensive drugs and bone health. Tinetti et al. linked antihypertensive medication with an elevated risk of severe fall-related injuries, particularly among those with a fall history^21^. Other studies have suggested that some antihypertensive drug classes, such as ACE inhibitors^22^, calcium channel blockers^22^, and renin-angiotensin system inhibitors^23^, might impact bone health. Notably, thiazide diuretics have been associated with lower fracture risk in some studies^24^. Nonetheless, a meta-analysis found no definitive evidence linking antihypertensive treatment with fracture risk^25^. As observational drug studies are susceptible to confounding by indication, comorbidity burden, treatment duration, and medication adherence, genetic approaches that proxy antihypertensive drug-target perturbation may provide complementary evidence for identifying blood pressure-lowering targets with skeletal relevance.

In this study, we leveraged individual-level data from the UK Biobank to investigate the relationship between blood pressure components and heel estimated bone mineral density (eBMD). We characterized linear and nonlinear associations of systolic and diastolic blood pressure with eBMD, examined DBP-stratified biomarker patterns, and evaluated genetically proxied antihypertensive-related target-gene expression in relation to eBMD using drug-target Mendelian randomization.

## Methods

### UK Biobank Data Sources and Statistical Analysis

The UK Biobank is a large-scale, long-term prospective cohort study that integrates extensive phenotypic, health-related, and genetic data. It recruited approximately 500,000 participants, collecting a broad array of phenotypic traits and biological samples. Genotyping was performed using the UK Biobank Axiom Array, and genotype imputation was conducted based on the 1000 Genomes Project Phase 3 reference panel^26^.

In this study, we utilized individual-level data from the UK Biobank (Application ID: 41376), consistent with the previous work^27–30^. To reduce population heterogeneity and potential confounding by ancestry, we restricted our analysis to participants of European descent, as inferred from genetic data. Genetic ancestry was inferred using the Rye algorithm, which estimates ancestry by performing principal component analysis (PCA) on reference genotypes^31^. Reference populations for PCA were obtained from the 1000 Genomes Project Phase 3, comprising 2,504 individuals across five continental ancestries: African (AFR), East Asian (EAS), European (EUR), South Asian (SAS), and Admixed American (AMR). For principal component computation, we merged UK Biobank data with the 1000 Genomes Project dataset using PLINK. A total of 565,631 HapMap3 variants were extracted following LD pruning (window size = 1000 kb, step size = 100, r^2^ = 0.9, minor allele frequency ≥ 0.01). After applying this ancestry inference and filtering for outliers, 366,443 participants remained for analysis.

The outcome phenotype in this study was heel bone mineral density (eBMD), which was derived from quantitative ultrasound (QUS) measurements using the formula: heel estimated BMD = 0.0025926 × (bone ultrasound attenuation + speed of sound) - 3.687 (Field ID: 3084, 3148, and 4105). Exposure variables included systolic and diastolic blood pressure (Field ID: 4080 and 4079). Additional covariates encompassed age at recruitment (Field ID: 21022), sex (Field ID: 31), assessment center (Field ID: 53), body mass index (Field ID: 21001), smoking status (Field ID: 20116), alcohol intake (Field ID: 20117), education (Field ID: 6138), and physical activity (Field ID: 884, 894, 904, 914). Physical activity was defined as adequate if participants met at least one of the following criteria: (1) moderate activity ≥5 days/week, (2) vigorous activity ≥1 day/week, (3) ≥150 minutes of moderate activity/week, or (4) ≥75 minutes of vigorous activity/week. We also included information on relevant comorbidities and medication usage. Disease diagnoses were obtained from ICD-9 and ICD-10 codes (Field ID: 41203 and 41270), as well as self-reported health conditions (Field ID: 20002). Medication usage data were derived from Field ID 20003. Baseline characteristics of the study population were provided in **Table S2.**

For statistical analysis, both linear and nonlinear regression models were applied. The “lm” function in R was used for linear regression, and the “ols” function from the “rms” package (R version 4.3.1) was employed to model nonlinear relationships via restricted cubic splines. Restricted cubic spline models were fitted with five knots, and nonlinearity was evaluated by testing the nonlinear spline terms. All observational models were adjusted for age at recruitment, sex, assessment center, BMI, smoking status, alcohol intake, education level, physical activity, diabetes diagnosis, antihypertensive medication use, glucocorticoid use, fracture history prior to baseline, and relevant comorbidities.

### Biomarker Stratified Analysis Based on Diastolic Blood Pressure

We stratified participants into two groups based on the median value of DBP. Individuals with DBP below the median were assigned to the Lower DBP Group, and those with DBP equal to or above the median were assigned to the Higher DBP Group.

Biomarker data were extracted from the baseline assessment of UK Biobank participants and included a broad panel of circulating and urinary markers reflecting liver and kidney function, lipid and glucose metabolism, hormonal activity, inflammation, and nutritional status.

Liver function was assessed using alanine aminotransferase (ALT, Field ID: 30620), aspartate aminotransferase (AST, Field ID: 30650), alkaline phosphatase (ALP, Field ID: 30610), gamma-glutamyl transferase (GGT, Field ID: 30730), total bilirubin (Field ID: 30840), direct bilirubin (Field ID:30660), albumin (Field ID: 30600), and total protein (Field ID: 30860). Kidney function was evaluated with serum creatinine (Field ID: 30700), cystatin C (Field ID: 30720), urea (Field ID: 30670), and urate (Field ID: 30880).

Lipid metabolism markers included total cholesterol (Field ID: 30690), HDL cholesterol (Field ID: 30760), LDL direct (Field ID: 30780), triglycerides (Field ID: 30870), apolipoprotein A (Field ID: 30630), apolipoprotein B (Field ID: 30640), and lipoprotein(a) (Field ID: 30790). Glucose metabolism was assessed using fasting glucose (Field ID: 30740) and glycated hemoglobin (HbA1c, Field ID: 30750). Hormonal markers included testosterone (Field ID: 30850), oestradiol (Field ID: 30800), insulin-like growth factor 1 (IGF-1, Field ID: 30770), and sex hormone-binding globulin (SHBG, Field ID: 30830). Inflammatory status was evaluated using C-reactive protein (CRP, Field ID: 30710) and rheumatoid factor (Field ID: 30820). Nutritional and mineral indicators included calcium (Field ID: 30680), phosphate (Field ID: 30810), and vitamin D (Field ID: 30890). Urinary biomarkers included urine creatinine (Field ID: 30510), sodium (Field ID: 30530), potassium (Field ID: 30520), and microalbumin (Field ID: 30500).

Extreme values, defined as observations exceeding three standard deviations from the mean, were excluded. Baseline characteristics of the study population were provided in **Table S4.**

Linear regression analyses were performed separately within the lower and higher DBP subgroups, defined by the median DBP value. All models were adjusted for age at recruitment, sex, assessment center, BMI, smoking status, alcohol intake, education level, physical activity, diabetes diagnosis, antihypertensive medication use, glucocorticoid use, fracture history prior to baseline, and relevant comorbidities, as described before.

To correct for multiple comparisons, Bonferroni correction was applied, with statistical significance defined as *P* < 0.05 divided by the number of biomarkers tested (*P* < 0.05/34= 1.47×10^-3^). Biomarkers were considered DBP-dependent if they exhibited opposite directions of association in the two subgroups and met the Bonferroni-corrected significance threshold in both groups.

### Drug-target Mendelian randomization of antihypertensive-related targets on eBMD

To estimate the potential causal relevance of genetically proxied antihypertensive-related target-gene expression to BMD, we performed a drug-target Mendelian randomization (MR) analysis^32,33^ using estimated bone mineral density (eBMD) as the outcome. The eBMD GWAS summary statistics were obtained from Morris et al^34^, which included 426,824 individuals of predominantly European ancestry and estimated eBMD from heel quantitative ultrasound measurements. An overview of the study design, including drug class selection, target-gene identification, eQTL instrument selection and Mendelian randomization framework, is presented in **Figure 1**.

**Figure 1.**
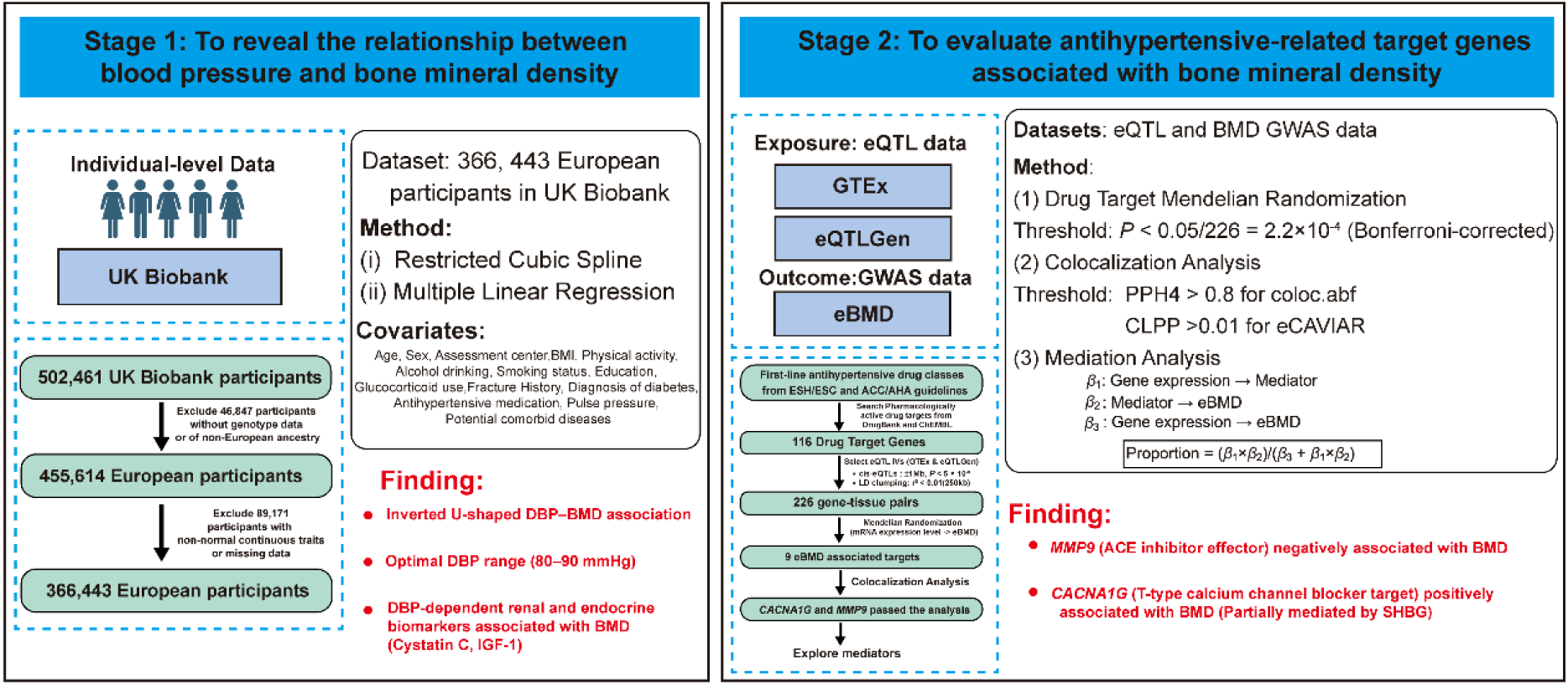
Study design. **Stage 1:** To evaluate the associations between blood pressure components (DBP and SBP) and estimated bone mineral density (eBMD) in 366,443 European-ancestry UK Biobank participants using restricted cubic splines and multivariable regression models. **Stage 2**: To identify first-line antihypertensive drug targets associated with eBMD by conducting drug-target Mendelian randomization with cis-eQTL instruments from GTEx and eQTLGen and eBMD GWAS data, followed by colocalization analyses (coloc.abf and eCAVIAR) and mediation analysis.

We first identified first-line antihypertensive drug classes based on international clinical guidelines from the ESH/ESC^35–37^ and ACC/AHA^38^. Active compounds within each class were retrieved, and their corresponding protein-coding target genes were obtained using the DrugBank and ChEMBL databases. In total, 116 drug target genes were included in the analysis.

To proxy target-gene expression, we used cis-expression quantitative trait loci (cis-eQTLs) as genetic instruments. eQTL data were derived from two sources. eQTLs across ten tissues relevant to cardiovascular and musculoskeletal systems were included from GTEx v8 dataset, including subcutaneous adipose tissue, visceral adipose tissue, aorta, coronary artery, tibial artery, cultured fibroblasts, atrial appendage, left ventricle, liver, and skeletal muscle^39^. Whole-blood eQTLs from both GTEx^39^ and eQTLGen^40^ were included to complement the selected tissue-specific eQTL datasets.

For each gene-tissue pair, we selected cis-eQTLs located within ±1 Mb of the gene region that reached genome-wide significance (*P* < 5 × 10^-8^). Independent SNPs were identified using PLINK (v1.9) with parameters --clump-kb 250 and --clump-r^2^ 0.01. MR estimates were obtained using the Wald ratio for gene–tissue pairs with a single instrument and IVW for those with multiple independent instruments in the MendelianRandomization package using R (version 4.3.1).

To account for potential confounding due to linkage disequilibrium (LD) or overlapping signals, we performed colocalization analyses using two complementary approaches. coloc.abf^41^ is an LD-independent Bayesian method that evaluates whether GWAS and eQTL signals are consistent with a shared causal variant. eCAVIAR^42^, in contrast, explicitly models LD structure and allows for the possibility of multiple causal variants. Default parameters were used for both methods. Evidence for colocalization was defined as posterior probability (PP.H4) > 0.8 for coloc.abf, or CLPP > 0.01 for eCAVIAR.

For genes that passed multiple-testing correction and colocalization analyses, we searched the Open Targets Genetics database (https://genetics.opentargets.org/)^43^ to identify associated phenotypes as candidate mediators. For each mediator, we first performed Mendelian randomization and colocalization analyses with gene expression as the exposure and the mediator as the outcome. When both analyses yielded positive results, we proceeded with Mendelian randomization using the mediator as the exposure and bone mineral density (BMD) as the outcome. Only those mediators that were positive in all tests were considered further.

To quantify the candidate indirect effect of the mediator on BMD, we used the product of coefficients method. The estimated indirect association through the candidate mediator was calculated as *β*_1_ × *β*_2_. The direct effect reflected the impact of gene expression on BMD, while the total effect combined both direct and indirect effects (*β*_3_ + *β*_1_ × *β*_2_). The proportion of the total effect mediated by the mediator was estimated by dividing the indirect effect by the total effect, i.e., [*β*_1_ ×*β*_2_/ (*β*_3_ + *β*_1_ × *β*_2_)].

## Results

### Inverted U-shaped DBP-eBMD association with renal and endocrine biomarker correlates

An overview of the study design was depicted in **Figure 1**. We first assessed the associations of blood pressure components, including diastolic blood pressure (DBP) and systolic blood pressure (SBP), with heel estimated bone mineral density (eBMD) among 366,443 European-ancestry UK Biobank participants (**Figure 1**). Baseline characteristics were shown in **Table S2**. Participants had a mean age of 56.72 years (SD 8.02) and mean eBMD of 0.54 (SD 0.12). Mean DBP and SBP were 81.88 (SD 10.33) mm Hg and 139.14 (SD 18.63) mm Hg, respectively. Men showed higher eBMD and higher DBP/SBP than women (all *P* < 0.001), and 22.4% of participants reported antihypertensive medication use. We observed a significant inverted U-shaped association between DBP and eBMD (*P*-nonlinear = 3.23 × 10^-9^). eBMD increased with DBP values below 80 mm Hg, reached higher levels within the 80-90 mm Hg range, and declined beyond 90 mm Hg (**Figure 2A, Table S3**). The pattern for SBP was similar to DBP, but nonlinear association was not significant for SBP (*P*-nonlinear = 0.18). Within the hypertensive range (DBP ≥ 90 mm Hg for DBP analysis; SBP ≥140 mm Hg for SBP analysis), both DBP and SBP were inversely associated with eBMD. The inverse association was numerically stronger for DBP (*β* = -3.36×10^-4^, SE =7.73×10^-5^, *P* =1.34×10^-5^) than for SBP (*β* = -8.05×10^-5^, SE = 3.38×10^-5^, *P* = 0.02) (**Figure 2A, Table S3**).

**Figure 2.**
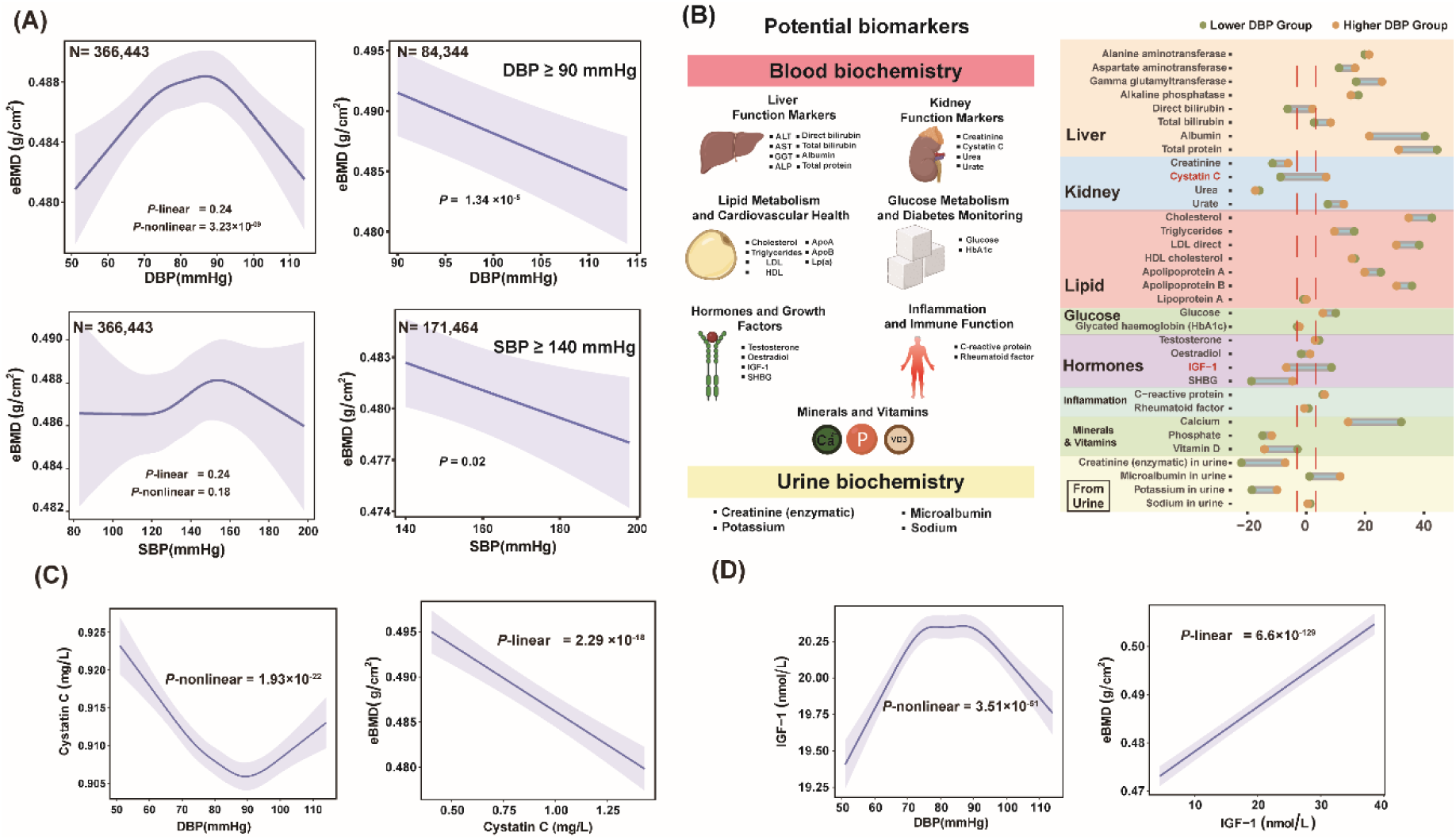
Inverted U-shaped DBP-eBMD relationship and DBP-dependent biomarker patterns. (A) Restricted cubic spline models for the nonlinear associations of DBP (top left) and SBP (bottom left) with eBMD in 366,443 UK Biobank European participants. Right panels show linear associations within the hypertensive ranges (DBP ≥ 90 mm Hg, N = 84,344; SBP ≥ 140 mm Hg, N = 171,464). (B) Exploration of DBP-dependent biomarker patterns related to eBMD. Left: Summary of blood and urine biomarkers categorized by physiological system. Right: Comparison of biomarker-eBMD associations between lower and higher DBP groups across multiple physiological categories. (C-D) Nonlinear (left) and linear (right) associations of DBP with cystatin C (C) and IGF-1 (D), and their respective associations with eBMD.

To explore biomarkers related to the nonlinear DBP-eBMD relationship, we examined 34 circulating and urinary biomarkers across liver, kidney, lipid, glucose, hormonal, inflammatory, mineral, and urinary domains (**Figure 2B left**). Most biomarkers showed similar directions of association with eBMD across the lower and higher DBP groups. However, cystatin C and IGF-1 exhibited distinct associations with eBMD depending on DBP levels (**Figure 2B right, Table S5**). We therefore focused on cystatin C and IGF-1 as DBP-dependent biomarker correlates of the nonlinear DBP-eBMD relationship. DBP showed a significant U-shaped association with cystatin C (*P*-nonlinear = 1.93 × 10^-22^), with lower levels observed at 90 mm Hg for DBP (**Figure 2C left**), while a strong negative linear association was observed between cystatin C and eBMD (*P*-linear = 2.29 × 10^-18^) (**Figure 2C right**). Furthermore, DBP exhibited an inverted U-shaped relationship with IGF-1 (*P*-nonlinear = 3.51 × 10^-51^), with peak concentrations at approximately 70-90 mm Hg for DBP (**Figure 2D left**), while IGF-1 levels were positively associated with eBMD in a linear manner (*P*-linear = 6.6 × 10^-129^) (**Figure 2D right**). These findings suggest that cystatin C and IGF-1 may help characterize renal and endocrine biomarker patterns related to the nonlinear association between DBP and eBMD.

### Drug-target MR prioritizes antihypertensive-related targets associated with eBMD

Using ESH/ESC and ACC/AHA guidelines together with DrugBank and ChEMBL, we identified 116 antihypertensive drug target genes (**Table S6**). We performed drug-target Mendelian randomization (MR) to prioritize antihypertensive-related genes with potential skeletal relevance. Genetically proxied target-gene expression was evaluated using eQTL data from 10 cardiovascular- and musculoskeletal-relevant GTEx tissues, together with whole-blood eQTL data from GTEx and eQTLGen (**Figure 1**). MR was performed only for gene-tissue pairs with valid cis-eQTL instruments, yielding 226 analyzable gene-tissue pairs after harmonization (**Table S7**). Notably, nine genes showed significant associations with eBMD after Bonferroni correction (*P* < 0.05/226 = 2.2 × 10^-4^). These included diuretic-related targets (*ALPL*, *SLC12A4*, and *SLC12A5*), calcium channel blocker-related targets (*CACNA2D3*, *CACNB3*, *CPT1A*, and *CACNA1G*), *ADRA2A* as an adrenergic neuron blocker-related target, and *MMP9* as an angiotensin-converting enzyme inhibitor-related target (**Figure 3A**, **Table S7**). Among these genes, *MMP9* and *CACNA1G* showed evidence of colocalization (CLPP= 0.06 for *MMP9*, PPH4.abf=0.97 for *CACNA1G*) (**Figure 3B**, **Table S8**). Specifically, *MMP9* was annotated as a captopril-related gene in drug-target databases (**Figure 3C**). Genetically proxied *MMP9* expression in cultured fibroblasts was negatively associated with eBMD (*β* = -0.036, *P* = 2.59 × 10^-6^). *CACNA1G* was annotated as a target of efonidipine and mibefradil, and genetically proxied *CACNA1G* expression in cultured fibroblasts was positively associated with eBMD (*β* = 0.042, *P* = 1.57 × 10^-9^) (**Figure 3D**, **Table S7**).

**Figure 3.**
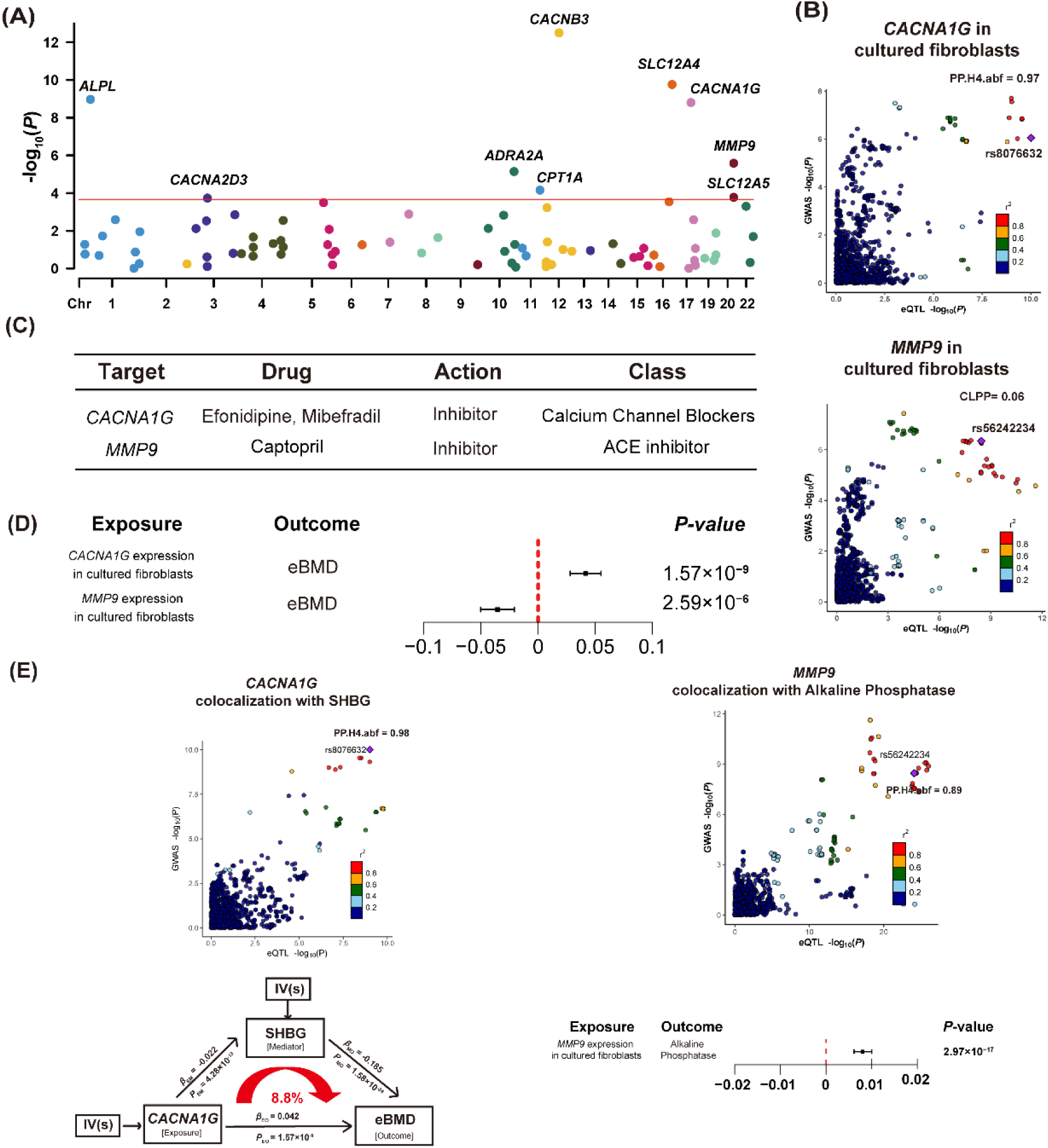
Drug-target MR prioritizes antihypertensive-related targets with potential relevance to eBMD. (A) Manhattan plot of MR results linking drug target gene expression to eBMD. Red solid line indicates *P* value threshold for significance (*P*< 2.2×10^-4^). Results are plotted by gene start position. (B) Colocalization plots for top gene–tissue pairs: *CACNA1G* and *MMP9* in cultured fibroblasts, showing evidence of colocalization between eQTL and eBMD GWAS signals. (C) Corresponding drugs and pharmacological classes for the identified targets. (D) Effect estimates and significance levels of prioritized gene–tissue pairs for eBMD. (E) Candidate intermediate phenotype analyses. SHBG was evaluated as a candidate mediator of the *CACNA1G*-eBMD association. Alkaline phosphatase was identified as a candidate phenotype associated with genetically proxied *MMP9* expression.

We next performed candidate mediation analyses to evaluate whether candidate intermediate phenotypes could partly account for the associations between prioritized genes and eBMD. For *MMP9*, genetically proxied expression was associated with alkaline phosphatase (*β*=7.99×10-3, *P*=2.97×10^-17^) (**Figure 3E right**, **Table S9**). For *CACNA1G*, SHBG was prioritized as a candidate mediator linking genetically proxied *CACNA1G* expression and eBMD (**Figure 3E left**, **Table S9**). *CACNA1G* expression was negatively associated with SHBG (*β* = -0.022, *P* = 4.28 × 10^-13^), and SHBG was inversely associated with eBMD (*β* = -0.185, *P* = 1.58×10^-24^). This yielded a positive candidate indirect pathway, with an estimated mediated proportion of 8.8%, while the direct association between *CACNA1G* expression and eBMD remained positive (*β* =0.042, *P* =1.57×10^−9^).

## Discussion

In this study, we observed a significant inverted U-shaped association between DBP and eBMD, with higher eBMD observed around a DBP range of 80-90 mm Hg. eBMD increased at lower DBP levels and declined in the hypertensive range, suggesting that the DBP-eBMD relationship may not be adequately captured by a simple linear model. Biomarker analyses indicated that cystatin C and IGF-1 showed DBP-dependent associations with eBMD, suggesting that renal and endocrine-related biomarkers may help characterize this nonlinear pattern. Drug-target Mendelian randomization prioritized genetically proxied *MMP9* and *CACNA1G* expression as antihypertensive-related signals associated with eBMD.

Both hypertension and osteoporosis are common aging-related diseases^44,45^. Previous studies have reported inconsistent associations between hypertension and BMD, with some showing positive associations between hypertension and BMD^12,13^, others suggesting no reduction in lumbar spine BMD among individuals with primary hypertension in non-Asian populations^46^, and several reporting negative associations ^47–49^. Our findings suggest that these discrepancies may partly reflect an underlying nonlinear relationship between DBP and bone density. Specifically, both lower and higher DBP ranges were associated with lower eBMD, whereas intermediate DBP values were associated with relatively higher eBMD. These results highlight the complexity of the blood pressure–bone density relationship and the importance of considering nonlinear patterns in future studies.

Our biomarker analyses provide further insight into this nonlinear relationship. Cystatin C and IGF-1 showed DBP-dependent associations with eBMD, suggesting that renal and endocrine-related biomarker patterns may help characterize the nonlinear association between DBP and eBMD. Elevated cystatin C may reflect impaired renal function and reduced acid excretion capacity. Impaired renal acid excretion may promote chronic low-grade metabolic acidosis, which can be buffered in part by bone, thereby promoting bone mineral dissolution, calcium mobilization, and osteoclast-mediated resorption^50–52^. As IGF-1 is a key anabolic factor that directly regulates osteoblast activity, bone formation, and bone mineral density^53,54^, reduced IGF-1 signaling may contribute to lower bone formation. Taken together, these findings indicate that renal and endocrine biomarkers, particularly cystatin C and IGF-1, may partly reflect the nonlinear association between DBP and eBMD.

The potential skeletal effects of antihypertensive pharmacotherapy have become an important area of investigation in aging populations. Large-scale population studies and causal inference approaches such as Mendelian randomization may provide complementary evidence for clarifying the inconsistent associations between antihypertensive treatment and skeletal outcomes^21–25^. In our drug-target MR analysis, genetically proxied MMP9 expression was associated with lower eBMD, consistent with the established role of MMP9 in osteoclastogenesis and extracellular matrix degradation^55^. *MMP9* was linked to captopril, an angiotensin-converting enzyme (ACE) inhibitor, in drug-target databases. Mechanistically, the local renin-angiotensin system in bone has been implicated in osteoclast activity, and angiotensin II may upregulate *MMP9* through AT1 receptor signaling^56,57^. This interpretation is consistent with preclinical studies showing that captopril attenuates bone loss and increases BMD in ovariectomized and hypertensive animal models^58–60^. However, observational studies of ACE inhibitors and skeletal health remain inconsistent, with reports of beneficial^61^, harmful^62^, or null associations^63^. These discrepancies may reflect differences between experimental and clinical contexts, including timing and duration of ACE inhibition, dose titration, treatment discontinuation, medication adherence, and comorbidity burden. In addition, the downstream effects of ACE inhibition may be complex, including bradykinin-related inflammatory pathways^64–66^, which could influence osteoclastogenesis^67,68^. Taken together, these findings support the potential skeletal relevance of MMP9 within antihypertensive pathways, while further studies are needed to clarify whether ACE inhibitor-related modulation of MMP9 influences bone outcomes.

Genetically proxied *CACNA1G* expression was positively associated with eBMD, suggesting that the *CACNA1G*-encoded T-type calcium channel may also have skeletal relevance. Some epidemiological studies have associated calcium channel blocker use with increased risks of osteoporosis or fracture^69^, but these findings require cautious interpretation because calcium channel blockers include drugs with different channel selectivity and clinical indications. Our mediation analysis further suggested that SHBG may partly account for the *CACNA1G-*eBMD association.

Genetically proxied *CACNA1G* expression was associated with lower SHBG, whereas SHBG was inversely associated with eBMD. Given that elevated SHBG may reduce the bioavailability of anabolic sex steroids^70,71^, this pathway may provide one possible explanation for the skeletal association of *CACNA1G*. Together with previous evidence suggesting impaired osteoblast function after T-type calcium channel blockade^69,72^, these findings highlight the need for further studies to evaluate T-type calcium channel-related pathways in bone health.

Several limitations should be acknowledged. First, the observational analyses were restricted to individuals of European ancestry, which may limit generalizability to other populations. Second, the observational analyses were based on baseline measurements, and residual confounding or reverse causation cannot be fully excluded. Third, eBMD was estimated from heel quantitative ultrasound rather than DXA-derived BMD, and fracture outcomes were not directly assessed. Fourth, the DBP-stratified biomarker analyses were exploratory and cannot establish mediation or causal mechanisms. Fifth, genetically proxied target-gene expression reflects lifelong genetic exposure and may not fully recapitulate pharmacological interventions, including drug-specific mechanisms, direction of target modulation, dose, timing, tissue specificity, treatment duration, and compensatory biological responses.

In conclusion, our study identified a nonlinear association between DBP and eBMD, with higher eBMD observed around a DBP range of 80–90 mm Hg.

Drug-target Mendelian randomization prioritized *MMP9*, an ACE inhibitor-related gene, and *CACNA1G*, a T-type calcium channel blocker target, as antihypertensive-related targets associated with eBMD. These findings suggest that antihypertensive-related target biology, including targets linked to captopril, efonidipine, and mibefradil, may have skeletal relevance.

### Perspectives

Skeletal health should be considered an important dimension of hypertension research, particularly in aging populations in whom blood pressure dysregulation and bone fragility frequently coexist. Future studies should assess bone density and fracture-related outcomes across blood pressure ranges, with attention to potential nonlinear associations. In parallel, longitudinal and experimental work is needed to clarify the skeletal relevance of antihypertensive-related target biology, particularly targets involving *MMP9* and *CACNA1G*.

### CRediT authorship contribution statement

JXG conceptualized the study, curated the data, performed formal analysis, conducted investigation, developed methodology, created visualizations, and wrote the original draft; MYY, XL, PW, ZHG, MYH, JSY, WJC, ZRL, SRG, JDZ, PPZ, BZ, ZHF, CLC, and DK contributed to validation, visualization, and writing – review & editing; HFZ conceptualized the study, supervised and managed the project, and critically reviewed and edited the manuscript. All authors read and approved the final manuscript.

## Acknowledgements

We would like to express our gratitude to the UK Biobank database, GTEx Consortium, the eQTLGen Consortium and the GEnetic Factors for OSteoporosis (GEFOS) Consortium for their valuable contributions. We also thank the Westlake University Supercomputer Centre for the facility support and technical assistance. This work was supported by Beijing GuoKe Biotechnology Co., LTD (Beijing, China) and China National GeneBank (CNGB) (Shenzhen, China). This work was supported by the National Key R&D Programme of China (#2024YFC3405703), and by the National Natural Science Foundation of China (#82370887).

## Data Availability Statement

The data supporting the findings of this study are available from publicly accessible repositories or upon reasonable request. Individual-level phenotypic and genetic data were obtained from the UK Biobank under Application ID 41376. GWAS summary statistics for heel estimated bone mineral density (eBMD) were derived from the GEFOS consortium (Morris et al.). Tissue-specific and whole blood eQTL data were obtained from the GTEx consortium (v8) and the eQTLGen consortium. Information regarding antihypertensive drug targets was sourced from the DrugBank and ChEMBL databases. All statistical analyses were performed using R (version 4.3.1), and custom scripts are available from the corresponding author upon reasonable request.

